# Transmission clusters, predominantly associated with men who have sex with men, play a main role in the propagation of HIV-1 in Northern Spain (2013-2018)

**DOI:** 10.1101/2021.09.28.21264185

**Authors:** Horacio Gil, Elena Delgado, Sonia Benito, Leonidas Georgalis, Vanessa Montero, Mónica Sánchez, Javier E. Cañada-García, Elena García-Bodas, Asunción Díaz, Michael M. Thomson, the Spanish Group for the Study of New HIV Diagnoses

**Affiliations:** HIV Biology and Variability Unit. Centro Nacional de Microbiología, Instituto de Salud Carlos III, Majadahonda, Madrid, Spain; HIV Surveillance and Behavioral Monitoring Unit. Centro Nacional de Epidemiología, Instituto de Salud Carlos III, Madrid, Spain

**Author notes:** These authors have contributed equally to this work and share first authorship (Horacio Gil, Elena Delgado). **Corresponding authors:** Michael M. Thomson) Horacio Gil), HIV Biology and Variability Unit, Centro Nacional de Microbiología. Instituto de Salud Carlos III, 28220 Majadahonda, Madrid, Spain.

**Keywords:** Spain, HIV-1, transmission clusters, molecular epidemiology, men who have sex with men, migrants

## Abstract

Viruses of HIV-1-infected individuals whose transmission is related group phylogenetically in transmission clusters (TCs). The study of the phylogenetic relations of these viruses and the factors associated with these individuals is essential to analyze the HIV-1 epidemic. In this study, we examine the role of TCs in the epidemiology of HIV-1 infection in Galicia and the Basque County, two regions of Northern Spain. A total of 1158 newly HIV-1-diagnosed patients (NDs) from both regions diagnosed in 2013-2018 were included in the study. Partial HIV-1 *pol* sequences were analyzed phylogenetically by approximately-maximum-likelihood with FastTree 2. In this analysis, 10,687 additional sequences from samples from HIV-1-infected individuals collected in Spain in 1999-2019 were also included to assign and determine the TCs’ sizes. TCs were defined as those which included viruses from ≥4 individuals, at least 50% of them Spaniards, and with ≥0.95 Shimodaira-Hasegawa-like node support in the phylogenetic tree. Factors associated to TCs were evaluated using odds ratios (OR) and their 95% confidence intervals (CI). Fifty one percent of NDs grouped in 162 TCs. Male patients (OR: 2.6; 95% CI: 1.5-4.7) and men having sex with men (MSM) (OR: 2.1; 95% CI: 1.4-3.2) had higher odds of belonging to a TC compared to female and heterosexual patients, respectively. Individuals from Latin America (OR: 0.3; 95% CI: 0.2-0.4), North Africa (OR: 0.4; 95% CI: 0.2-1.0), and especially Sub-Saharan Africa (OR: 0.02; 95% CI: 0.003-0.2) were inversely associated to belonging to TCs compared to native Spaniards. Differences in distribution and sizes of local TCs were observed in both regions reflecting a different epidemic pattern. Our results show that TCs play an important role in the spread of HIV-1 infection in the two Spanish regions studied, where transmission between MSM is predominant. The majority of migrants were infected with viruses not belonging to TCs that expand in Spain. Molecular epidemiology is essential to identify local peculiarities of HIV-1 propagation. The early detection of TCs and prevention of their expansion, implementing effective control measures, could reduce HIV-1 infections.

## INTRODUCCION

The HIV-1 epidemic is still one of the major public health problems in Spain. Around 3,500 - 4,000 new diagnoses of HIV-1 infection (NDs) are reported every year, with an estimated incidence of NDs of 7.5 per 100,000 population in 2019 (HIV, STI and hepatitis surveillance unit, 2020). A decreasing trend in the incidence of NDs has been observed since 2010, although it is still higher than the average rate found in the EU/EEA (European Centre for Disease Prevention and Control and WHO Regional Office for Europe, 2020). In 2019, most (56%) of the NDs were diagnosed among men who had sex with men (MSM) and 36% of all reported NDs were in people born outside of Spain (HIV, STI and hepatitis surveillance unit, 2020).

Molecular epidemiology is an important tool for describing the HIV-1 epidemic (Board et al., 2020; Brenner et al., 2013; Paraskevis et al., 2016; Wertheim et al., 2017). Individuals whose transmission is related group phylogenetically in clades named transmission clusters (TCs). Due to the high genetic variability of HIV-1, phylogenetic analysis allows reconstructing transmission events through the identified TCs and infer the history of the HIV-1 epidemic (Hué et al., 2004; Hué et al., 2005).

National databases of protease and reverse transcriptase (Pr-RT) sequences, primarily obtained for antiretroviral drug resistance testing, contain valuable data about HIV-1 expansion and have been used in molecular epidemiology studies (Fabeni et al., 2021; Oster et al., 2018; Parczewski et al., 2017; Petersen et al., 2018; Pineda-Peña et al., 2019; Verhofstede et al., 2018). The phylogenetic analyses performed in these studies combined with clinical and epidemiological data of the patients provide relevant public health information for the implementation of control measures and for monitoring the HIV-1 epidemic (Board et al., 2020; Brenner et al., 2013; Campbell et al., 2020; Paraskevis et al., 2019; Vasylyeva et al., 2016; Vasylyeva et al., 2020).

Spanish clinical guidelines recommend to perform a genotypic drug resistance test before starting antiretroviral therapy (ART) in all HIV-1-diagnosed patients and in ART-failing cases (AIDS study group (GESIDA) of the Spanish Society of Infectious Diseases and Clinical Microbiology and the national AIDS Plan., 2020). Pr-RT sequences obtained for these tests have been analyzed in different molecular epidemiology studies, describing the genetic features of the HIV-1 epidemic in different regions of Spain (Cuevas et al., 2009a; Cuevas et al., 2009b; González-Alba et al., 2011; González-Domenech et al., 2020; Holguín et al., 2007; Patiño-Galindo et al., 2016; Patiño-Galindo et al., 2017b; Pérez-Parra et al., 2016; Pérez-Parra et al., 2015; Thomson et al., 2001; Yebra et al., 2013). The phylogenetic studies have also allowed the identification of large TCs among MSM which are actively growing in Spain (Delgado et al., 2015; Delgado et al., 2019; González-Domenech et al., 2018; Patiño-Galindo et al., 2017a), as well as transmitted drug resistance mutations which are spreading in TCs (Cuevas et al., 2009b; González-Domenech et al., 2018; González-Domenech et al., 2020; Vega et al., 2015; Viciana et al., 2016), highlighting the value of molecular epidemiology as a tool for HIV surveillance.

The Basque Country and Galicia are two regions located in northern Spain, comparable in terms of population and HIV-1 diagnose rates. During 2013-2018 the mean resident population and rates of new HIV diagnoses were estimated in 2,2 million and 6,9 per 100,000 inhabitants for the Basque Country and 2,7 million and 5,6 per 100,000 inhabitants for Galicia (HIV, STI and hepatitis surveillance unit, 2020; Instituto Nacional de Estadistica, 2021). In this study, we analyze the role of TCs in the transmission of HIV-1 infection among patients diagnosed in 2013-2018 in these northern Spanish regions, identifying the different features associated with TCs in the HIV-1 epidemics in these regions.

## MATERIAL AND METHODS

### Patients

Samples from individuals newly diagnosed of HIV-1 infection in the northern Spanish regions of Galicia and Basque Country during 2013-2018, sent to the HIV Biology and Variability Unit, Centro Nacional de Microbiología, Instituto de Salud Carlos III, were included in the study.

The representativeness of the NDs of our cohort was estimated comparing them to the NDs reported to the Spanish information system on new HIV diagnoses (SINIVIH by its Spanish acronym)(HIV, STI and hepatitis surveillance unit, 2020), from the same period and studied regions. As patients in both databases cannot be linked, gender, age group, transmission route and region of birth frequencies were compared between both groups of individuals using the chi-squared test.

### Nucleic acid extraction, amplification and sequencing

Nucleic acid was extracted from plasma or whole blood samples. RNA was extracted from 1 ml plasma using NUCLISENS® easyMAG® (BioMérieux, Marcy l’Etoile, France) and DNA was extracted from 200 µl whole blood using QIAamp® DNA DSP blood mini kit (Qiagen, Hilden, Germany), following the manufacturer’s instructions. A Pr-RT fragment of *pol* (HXB2 positions 2253-3629) was amplified by RT-PCR followed by nested PCR from RNA or by nested PCR from DNA. Reagents, PCR thermal profiles, and primers are described in supplementary tables 1 and 2.

Population sequencing was performed with ABI Prism BigDye Terminator Cycle Sequencing kit and ABI 3730 XL sequencer (Applied Biosystems, Foster City, CA, U.S.A.) in the Genomic Unit of Instituto de Salud Carlos III. Sequences were assembled with SeqMan Pro v.12.2.1 (DNA STAR Lasergene, Madison, WI, USA) and edited with BioEdit v.7.2.5 (Hall, 1999) (http://www.mbio.ncsu.edu).

### Phylogenetic analyses and transmission cluster identification

Partial *pol* sequences from the patients included in this study and sequences obtained by us from another 10,687 HIV-1-infected individuals attended in clinical centers from Spain whose samples were collected in 1999-2019 were included in the analyses. Sequences were analyzed phylogenetically by an approximately maximum-likelihood method using FastTree2 (Price et al., 2010). In these analyses, the general time reversible model of nucleotide substitution with CAT approximation to account for among-site heterogeneity in substitution rates was used, and the reliability of nodes was assessed with Shimodaira-Hasegawa(SH)-like local support values. Classification of sequences in subtypes and recombinant forms was based on clustering with clade references in approximately maximum-likelihood trees. Sequences suspected of intersubtype recombination, were subsequently analyzed by bootscanning with SimPlot v3.5 (Lole et al., 1999)

TCs were defined as those comprising viruses from four or more individuals, at least 50% of them Spanish, and whose sequences grouped in the phylogenetic tree with a SH-like node support value ≥0.95.

### Statistical analysis

To evaluate the association between variables, chi-squared and Fisher’s exact tests were used, with associations being considered statistically significant at a *p*-value <0.05. A multivariable logistic regression model was performed to identify factors associated to TCs. The model was adjusted by sex, age group, transmission route, country of origin of the patient, Spanish region of sample collection, and HIV-1 genetic form. Associations were measured using the odds ratio (OR) and its 95% confidence interval (CI). Data analyses were performed using the STATA statistical software package Version 16 (Stata Corporation, College Station, TX, US).

## RESULTS

### Study Population

A total of 1,158 HIV-1 NDs from Galicia and Basque Country diagnosed in 2013-2018 were included in the study (Table 1). The majority of the patients were male (82%), with MSM being the most common transmission route (46%). Heterosexuals represented 34%, and 15% of the individuals were male whose transmission route was reported as sexual without specifying whether they were in the MSM or heterosexual category. Subtype B (63%) was the most frequent genetic form among the individuals included in the study, followed by unique recombinant forms (URFs) (8.2%), CRF02_AG (7.7%), and subtype F (7.6%) (Table 1).

**Table 1.** Characteristics of the HIV-1 newly diagnosed individuals included in the study.

|  |  | BC* |  | GA† |  | All |  | p-value |
| --- | --- | --- | --- | --- | --- | --- | --- | --- |
|  |  | N | % | N | % | N | % |  |
| Gender | Male | 626 | 82 | 306 | 83 | 932 | 82 | 0.71 |
|  | Female | 133 | 17 | 65 | 17 | 198 | 17 |  |
|  | Transexual | 5 | 0.65 | 0 | 0 | 5 | 0.44 |  |
|  | Unknown | 12 |  | 11 |  | 23 |  |  |
| Transmission route | MSM ‡ | 317 | 47 | 142 | 44 | 459 | 46 | 0.61 |
|  | Heterosexual | 233 | 34 | 112 | 35 | 345 | 34 |  |
|  | MNSST § | 103 | 15 | 48 | 15 | 151 | 15 |  |
|  | PWID ¶ | 22 | 3.2 | 17 | 5.3 | 39 | 3.9 |  |
|  | Other | 6 | 0.88 | 3 | 0.93 | 9 | 0.90 |  |
|  | Unknown | 95 |  | 60 |  | 155 |  |  |
| Region of origin | Spain | 477 | 65 | 268 | 83 | 745 | 70 | <0.001 |
|  | Latin America | 147 | 20 | 37 | 11 | 184 | 17 |  |
|  | Sub-Saharan Africa | 79 | 11 | 6 | 1.9 | 85 | 8.0 |  |
|  | North Africa | 14 | 1.9 | 1 | 0.3 | 15 | 1.4 |  |
|  | Europe | 16 | 2.2 | 12 | 3.7 | 28 | 2.6 |  |
|  | Other | 5 | 0.68 | 1 | 0.31 | 6 | 0.56 |  |
|  | Unknown | 38 |  | 57 |  | 95 |  |  |
| Genetic form | B | 489 | 63 | 235 | 62 | 724 | 63 | <0.001 |
|  | F | 39 | 5.0 | 49 | 13 | 88 | 7.6 |  |
|  | A | 21 | 2.7 | 12 | 3.1 | 33 | 2.9 |  |
|  | C | 19 | 2.5 | 20 | 5.2 | 39 | 3.4 |  |
|  | G | 12 | 1.6 | 8 | 2.1 | 20 | 1.7 |  |
|  | CRF02_AG | 77 | 9.9 | 12 | 3.1 | 89 | 7.7 |  |
|  | CRF_BF | 20 | 2.6 | 8 | 2.1 | 28 | 2.4 |  |
|  | URF | 65 | 8.4 | 30 | 7.9 | 95 | 8.2 |  |
|  | Other | 34 | 4.4 | 8 | 2.1 | 42 | 3.6 |  |
| Total |  | 776 |  | 382 |  | 1158 |  |  |

\*BC: Basque Country; †GA: Galicia; ‡MSM: Men who have sex with men; §MNSST: Men who have non-specified sexual transmission; ¶PWID: Person who injects drugs.

Seventy percent of the patients were native Spanish (Table 1), followed by Latin Americans (17%) and Sub-Saharan Africans (8.0%). There were important differences in gender and transmission route proportions between these two migrant populations, being higher in Sub-Saharan Africans than in Latin Americans: female [52% (44/85) vs. 26% (37/180)] and heterosexual transmission [88% (65/74) vs. 33%. (54/165)], respectively.

The distribution of patients by gender and transmission route was similar in both studied regions (Table 1), although the frequency of people who inject drugs (PWID) was slightly higher in Galicia (5.3% vs. 3.2%). The percentage of patients of non-Spanish origin was 35% in the Basque Country (13% corresponding to Africans), a proportion which is statistically higher (*p*<0.001) compared with the 17% found in Galicia (2.2% Africans). Statistical differences in the distribution of genetic forms were also observed between both regions (*p*<0.001), with the frequencies of subtype F (13%) and CRF02_AG (9.9%) being higher in Galicia and the Basque Country, respectively (Table 1).

### Representativeness of the patients included in the study

Our cohort of newly diagnosed HIV-1-infected patients represents 64% of the notified cases in 2013-2018 (Table 2), assuming that all NDs included in our study have been reported to the SINIVIH. This percentage was higher in the Basque Country, where it reached 84%, than in Galicia, where it was 41% of the notified cases. No statistical differences were found between both patient groups regarding the distribution of individuals by gender, transmission route, region of origin or age group (Table 2).

**Table 2.** New HIV-1 diagnoses from Basque Country and Galicia (2013-18) included in the study and reported to the HIV National Surveillance System.

|  |  | Laboratory |  | Surveillance* |  | *p*-value |
| --- | --- | --- | --- | --- | --- | --- |
|  |  | N† | % | N | % |  |
| Gender | Male | 932 | 82 | 1477 | 81 | 0.461 |
|  | Female | 198 | 18 | 340 | 19 |  |
|  | Unknown & Other‡ | 28 |  | 0 |  |  |
| Transmission route | MSM § | 459 | 54 | 957 | 57 | 0.096 |
|  | Heterosexual | 345 | 41 | 631 | 37 |  |
|  | PWID ¶ | 39 | 4.6 | 104 | 6.1 |  |
|  | MNSST ¶ | 151 |  | NA‡ |  |  |
|  | Unknown & Other‡ | 164 |  | 125 |  |  |
| Age group | <20 | 25 | 2.2 | 5 | 1.5 | 0.131 |
|  | 20-29 | 256 | 23 | 23 | 21 |  |
|  | 30-39 | 396 | 35 | 151 | 33 |  |
|  | ≥40 | 456 | 40 | 280 | 44 |  |
|  | Unknown‡ | 25 |  | 0 |  |  |
| Region of origin | Spain | 745 | 70 | 1259 | 70 | 0.881 |
|  | Latin America | 184 | 17 | 301 | 17 |  |
|  | Sub-Saharan Africa | 85 | 8.0 | 159 | 8.9 |  |
|  | North Africa | 15 | 1.4 | 22 | 1.2 |  |
|  | Europe | 28 | 2.6 | 54 | 3.0 |  |
|  | Unknown & Other‡ | 101 |  | 22 |  |  |
| Total | Both Regions | 1158 | 64 | 1817 | 100 |  |
|  | Basque Country | 759 | 84 | 907 | 100 |  |
|  | Galicia | 371 | 41 | 910 | 100 |  |

\* Data from the Spanish Information Systems on HIV New Diagnoses (SINIVIH); † N: number of patients; ‡ Sexual male, unknown and others categories were not included in the percentage and *p*-value calculations; § MSM: men who have sex with men; ¶ PWID: person who injects drugs; ¶ MNSST: Men who have non-specified sexual transmission; ‡ NA: Not applicable, MNSST is not a category collected by SINIVIH.

### TCs identified in the study

Fifty one percent (594/1158) of the individuals belonged to 162 different TCs. Regarding the regions, 45% (351/776) of the patients were included in 98 TCs in the Basque Country and 64% (243/382) belonged to 82 TCs in Galicia (Table 3). Eighteen (11%) TCs comprised patients from both regions. The sizes of the TCs ranged from 4 to 204 patients. MSM was the main transmission route associated with 59% of the TCs (Table 3). Transmission routes other than MSM were more frequently associated with TCs in Galicia (44%) than in the Basque County (35%) (Table 3), although the difference was not statistically significant (*p*=0.207).

**Table 3:** Characteristic of the identified TCs of the study.

|  |  | Basque Country |  | Galicia |  | Both regions |  |
| --- | --- | --- | --- | --- | --- | --- | --- |
|  |  | N | % | N | % | N | % |
| Transmission route* | MSM† | 64 | 65 | 46 | 56 | 95 | 59 |
|  | Heterosexual | 10 | 10 | 14 | 17 | 24 | 15 |
|  | Sexual‡ | 16 | 16 | 10 | 12 | 23 | 14 |
|  | PWID § | 8 | 8.2 | 12 | 15 | 20 | 12 |
| Total |  | 98 | 100 | 82 | 100 | 162 | 100 |

\* Main mode of HIV transmission among individuals in the TC; †MSM: men who have sex with men; ‡ There is not predominance of MSM or heterosexual transmission; § PWID: Persons who inject drugs.

A total of 15 large TCs (here defined as those with ≥30 individuals) were identified in the study (Table 4), 10 of them of subtype B. MSM was the associated transmission route in 14 (93%) of them. Most of these large TCs have spread mainly in a specific region. Thus, TCs B07, B08, B09, B10, B12, B70, and F1_3 were found mainly in the Basque Country, while TCs F1_1, B13, BG_2, and B05 were spread mainly in Galicia (Table 4). In addition, A1_1, CRF02_1, B50 and B31 have a global expansion in Spain, with a high number of patients outside of both studied regions. Four TCs had greater than 50% increase in NDs during the period 2013-2018: F1_3 (94%), A1_1 (76%), B70 (72%) and CRF02_1 (53%) (Table 4).

**Table 4.** Characteristics of the transmission clusters comprising ≥ 30 patients in the Basque Country and Galicia (2013-2018).

| Cluster | Genetic form | No. patients | Regional distribution |  |  |  |  |  | % Spanish | % Male | Main trasmission route | Patients (2013-18)‡ |  | Period§ |
| --- | --- | --- | --- | --- | --- | --- | --- | --- | --- | --- | --- | --- | --- | --- |
|  |  |  | BC* |  | GA† |  | Others |  |  |  |  | N | % |  |
|  |  |  | N | % | N | % | N | % |  |  |  |  |  |  |
| F1_1 | F1 | 205 | 9 | 4.4 | 138 | 67 | 58 | 28 | 85 | 97 | MSM | 78 | 38 | 2000-2019 |
| CRF02_1 | CRF02_AG | 83 | 34 | 41 | 3 | 3.6 | 46 | 55 | 79 | 92 | MSM | 44 | 53 | 2008-2019 |
| B50 | B | 74 | 15 | 20 | 3 | 4.1 | 56 | 76 | 85 | 100 | MSM | 34 | 46 | 2007-2019 |
| A1_1 | A1 | 63 | 8 | 13 | 24 | 38 | 31 | 49 | 71 | 95 | MSM | 48 | 76 | 2006-2019 |
| B70 | B | 61 | 56 | 92 |  |  | 5 | 8.2 | 88 | 98 | MSM | 44 | 72 | 2008-2019 |
| B08 | B | 60 | 46 | 77 | 6 | 10 | 8 | 13 | 92 | 98 | MSM | 13 | 22 | 2006-2019 |
| B09 | B | 57 | 55 | 97 |  |  | 2 | 3.5 | 88 | 98 | MSM | 10 | 18 | 1991-2019 |
| B13 | B | 55 |  |  | 54 | 98 | 1 | 1.8 | 89 | 94 | MSM | 16 | 29 | 2004-2017 |
| B31 | B | 49 | 19 | 39 | 2 | 4.1 | 28 | 57 | 77 | 98 | MSM | 23 | 47 | 2001-2019 |
| B07 | B | 38 | 36 | 95 |  |  | 2 | 5.3 | 91 | 95 | MSM | 7 | 18 | 2000-2019 |
| B10 | B | 36 | 29 | 81 |  |  | 7 | 19 | 90 | 97 | MSM | 7 | 19 | 1991-2019 |
| F1_3 | F1 | 36 | 29 | 81 |  |  | 7 | 19 | 76 | 100 | MSM | 34 | 94 | 2014-2018 |
| B12 | B | 35 | 25 | 71 | 1 | 2.9 | 9 | 26 | 63 | 100 | MSM | 11 | 31 | 2009-2019 |
| BG_2 | URF_BG | 33 |  |  | 33 | 100 |  |  | 85 | 80 | PWID | 2 | 6 | 1994-2019 |
| B05 | B | 31 |  |  | 31 | 100 |  |  | 93 | 94 | MSM | 10 | 32 | 2003-2019 |

1. \*BC: Basque Country; †GA: Galicia; ‡ Newly-diagnosed HIV-1 patients during 2013-2018; §Year of HIV-1 diagnosis of the patients in the TC.

### Factors associated with TCs

The percentage of patients in TCs was higher in males (57%) than in females (22%) and in MSM (67%) compared to heterosexuals (31%). This percentage was also higher in Spaniards (64%) than in Latin Americans (35%) or in Sub-Saharan Africans (1.2%) and in individuals infected with viruses of subtypes F (76%) or B (58%) compared to those infected with CRF02_AG viruses (17%). The different distribution of TCs according to gender, mode of HIV-1 transmission, country of origin of the patient and HIV-1 genetic form were statistically significant (Table 5). The age group was also associated to TCs (p*=*0.036). The age range of 20-29 years showed an increased percentage of patients in TCs (59%), which was especially high in Galicia (81%, p=0.003) but was not statistical significant in the Basque Country, and no statistical differences were found in the distribution of main variables

**Table 5:** Characteristics of the new HIV diagnoses included in TCs (2013-2018).

| Variables | Categories | Basque Country |  |  |  |  | Galicia |  |  |  |  | Both regions |  |  |  |  |
| --- | --- | --- | --- | --- | --- | --- | --- | --- | --- | --- | --- | --- | --- | --- | --- | --- |
|  |  | No |  | Yes |  | p-value | No |  | Yes |  | p-value | No |  | Yes |  | p-value |
|  |  | N* | % | N | % |  | N | % | N | % |  | N | % | N | % |  |
| Gender<br>(n= 1135, no data=23) | Female | 115 | 86 | 18 | 14 | <0.001 | 40 | 62 | 25 | 38 | <0.001 | 155 | 78 | 43 | 22 | <0.001 |
|  | Male | 303 | 48 | 323 | 52 |  | 95 | 31 | 211 | 69 |  | 398 | 43 | 534 | 57 |  |
|  | Transexual | 3 | 60 | 2 | 40 |  | 0 | 0 | 0 | 0 |  | 3 | 60 | 2 | 40 |  |
| Transmission route<br>(n=1003, no data=155) | Heterosexual | 178 | 76 | 55 | 24 | <0.001 | 60 | 54 | 52 | 46 | <0.001 | 238 | 69 | 107 | 31 | <0.001 |
|  | MSM† | 120 | 38 | 197 | 62 |  | 33 | 23 | 109 | 77 |  | 153 | 33 | 306 | 67 |  |
|  | MNSST‡ | 48 | 47 | 55 | 53 |  | 18 | 37 | 30 | 63 |  | 66 | 43 | 85 | 56 |  |
|  | PWID§ | 15 | 68 | 7 | 32 |  | 8 | 47 | 9 | 53 |  | 23 | 59 | 16 | 41 |  |
|  | Other | 4 | 67 | 2 | 33 |  | 3 | 100 | 0 | 0 |  | 7 | 78 | 2 | 22 |  |
| Region of origin (n=1063, no data=95) | Spain | 189 | 40 | 288 | 60 | <0.001 | 82 | 31 | 186 | 69 | 0.001 | 271 | 37 | 474 | 64 | <0.001 |
|  | Latin America | 102 | 69 | 45 | 31 |  | 18 | 49 | 19 | 51 |  | 120 | 65 | 64 | 35 |  |
|  | Sub-Saharan Africa | 79 | 100 | 0 | 0 |  | 5 | 83 | 1 | 17 |  | 84 | 99 | 1 | 1.2 |  |
|  | North Africa | 9 | 64 | 5 | 36 |  | 1 | 100 | 0 | 0 |  | 10 | 67 | 5 | 33 |  |
|  | Europe | 11 | 69 | 5 | 31 |  | 7 | 58 | 5 | 42 |  | 18 | 64 | 10 | 36 |  |
|  | Other | 4 | 80 | 1 | 20 |  | 1 | 100 | 0 | 0 |  | 5 | 83 | 1 | 17 |  |
| Age group<br>(n=1133, no data=25) | <20 | 10 | 53 | 9 | 47 | 0.44 | 3 | 50 | 3 | 50 | 0.003 | 13 | 52 | 12 | 48 | 0.036 |
|  | 20-29 | 92 | 51 | 90 | 49 |  | 14 | 19 | 60 | 81 |  | 106 | 41 | 150 | 59 |  |
|  | 30-39 | 160 | 58 | 115 | 41 |  | 49 | 41 | 72 | 59 |  | 209 | 53 | 187 | 47 |  |
|  | ≥40 | 161 | 55 | 137 | 45 |  | 68 | 43 | 90 | 57 |  | 229 | 50 | 227 | 50 |  |
| Genetic form<br>(n=1158, no data=0) | A | 14 | 67 | 7 | 33 | <0.001 | 3 | 25 | 9 | 75 | 0.002 | 17 | 52 | 16 | 48 | <0.001 |
|  | B | 224 | 46 | 265 | 54 |  | 83 | 35 | 152 | 65 |  | 307 | 42 | 417 | 58 |  |
|  | C | 11 | 58 | 8 | 42 |  | 8 | 40 | 12 | 60 |  | 19 | 49 | 20 | 51 |  |
|  | F | 10 | 26 | 29 | 74 |  | 11 | 23 | 38 | 78 |  | 21 | 24 | 67 | 76 |  |
|  | G | 10 | 83 | 2 | 17 |  | 5 | 63 | 3 | 37 |  | 15 | 75 | 5 | 25 |  |
|  | CRF02_AG | 63 | 82 | 14 | 18 |  | 11 | 92 | 1 | 8.3 |  | 74 | 83 | 15 | 17 |  |
|  | CRF_BF | 18 | 90 | 2 | 10 |  | 4 | 50 | 4 | 50 |  | 22 | 79 | 6 | 21 |  |
|  | URF | 47 | 72 | 18 | 28 |  | 10 | 33 | 20 | 67 |  | 57 | 60 | 38 | 40 |  |
|  | Other | 28 | 82 | 6 | 18 |  | 4 | 50 | 4 | 50 |  | 32 | 76 | 10 | 24 |  |

\*N: Number of patients; † MSM: men who have sex with men; ‡ MNSST: Men who have non-specified sexual transmission; §PWID: People who inject drugs.

In the multivariate analysis, males (OR: 2.6; 95% CI: 1.5-4.7) and MSM (OR: 2.1; 95% CI: 1.4-3.2) had higher odds of belonging to a TC compared to females and heterosexuals, respectively (Table 6). Patients from Galicia (OR: 1.5; 95% CI: 1.1-2.2) and subtype F infections (OR: 2.5; 95% CI: 1.3-5.0) were also factors associated with TCs (Table 6)

**Table 6:** Factors associated with transmission clusters, univariate/multivariate analysis.

| Variables |  | Univariate analysis |  | Multivariate analysis |  |
| --- | --- | --- | --- | --- | --- |
|  |  | OR* | 95% CI† | Adjusted OR | 95% CI |
| Gender | Female | Reference |  | Reference |  |
|  | Male | 4.8 | 3.3-7.1 | 2.6 | 1.5-4.7 |
|  | Transexual | 2.4 | 0.4-15 | NA‡ | NA |
| Transmission route | Heterosexual | Reference |  | Reference |  |
|  | MSM§ | 4.5 | 3.2-6.1 | 2.1 | 1.4-3.2 |
|  | MNNSST¶ | 2.9 | 1.9-4.3 | 1.4 | 0.8-2.3 |
|  | PWIDⓈ | 1.6 | 0.8-3.1 | NA | NA |
|  | Other | 0.6 | 0.1-3.1 | NA | NA |
| Region of origin | Spain | Reference |  | Reference |  |
|  | Latin America | 0.3 | 0.2-0.4 | 0.3 | 0.2-0.4 |
|  | Sub-Saharan Africa | <0.01 | <0.001-0.06 | 0.02 | 0.003-0.2 |
|  | North Africa | 0.3 | 0.1-0.9 | 0.4 | 0.2-1.0 |
|  | Europe | 0.3 | 0.1-0.7 | 0.3 | 0.03-3.5 |
|  | Other | 0.1 | 0.1-1.0 | 0.4 | 0.1-1.6 |
| Age group | <20 | 0.7 | 0.3-1.5 | NA | NA |
|  | 20-29 | Reference |  | Reference |  |
|  | 30-39 | 0.6 | 0.5-0.9 | 0.6 | 0.4-0.9 |
|  | ≥40 | 0.7 | 0.5-1.0 | 0.6 | 0.4-0.9 |
| Spanish Region | Basque Country | Reference |  | Reference |  |
|  | Galicia | 2.1 | 1.6-2.7 | 1.5 | 1.1-2.2 |
| Genetic form | A | 0.7 | 0.3-1.4 | NA | NA |
|  | B | Reference |  | Reference |  |
|  | C | 0.8 | 0.4-1.5 | NA | NA |
|  | F | 2.4 | 1.4-3.9 | 2.5 | 1.3-5.0 |
|  | G | 0.3 | 0.1-0.7 | 0.4 | 0.1-1.5 |
|  | CRF02_AG | 0.2 | 0.1-0.3 | 0.6 | 0.3-1.2 |
|  | CRF_BF | 0.2 | 0.1-0.5 | 0.1 | 0.03-0.4 |
|  | URF | 0.5 | 0.3-0.8 | 0.8 | 0.4-1.4 |
|  | Other | 0.4 | 0.3-0.7 | 0.3 | 0.2-0.8 |

\*OR: Odds ratio; † 95% CI: 95% confidence interval; ‡NA: Not applicable. Adjusted OR were not calculated for categories with $p < 0.1$ in the univariate analysis; § MSM: Men having sex with men; ¶ MNSST: Men who have non-specified sexual transmission; Ⓢ PWID: People who inject drugs.

Individuals from Latin America (OR: 0.3; 95% CI: 0.2-0.4), North Africa (OR: 0.4; 95% CI: 0.2-1-0) and especially from Sub-Saharan Africa (OR: 0.02; 95% CI: 0.003-0.2) were inversely associated to belonging to TCs compared to Spanish patients (Table 6). Similarly, patients over 29 years old (OR: 0.6; 95% CI: 0.4-0.9) and infections with a virus of a CRF_BF (OR: 0.1; 95% CI: 0.03-0.4) or of a genetic form classified in the “other” category (OR 0.3 95% CI: 0.2-0.8) had lower odds of belonging to TCs than patients in the age range of 20-29 years and with subtype B infections, respectively (Table 6).

## DISCUSSION

The phylogenetic analysis of almost 12,000 HIV-1-infected patients from our database has allowed us to investigate the transmission events occurring during the 2013-2018 period in two regions of Northern Spain, Galicia and Basque Country, identifying TCs and their associated factors. Our ND cohort had a good representativeness, corresponding to 64% of the NDs reported to the SINIVIH from these regions during that period, especially in the Basque Country where it reached 84%.

Our analysis has assigned 51% of the NDs from Galicia and Basque Country to 162 different TCs, indicating that TCs are playing an important role in the spread of HIV-1 infections in both regions. There is no consensus on the criteria for defining a TC, as these should depend on the aim of the study (Hassan et al., 2017). We considered only TCs including at least 4 documented infections. Therefore, our more strict TC definition, compared to some other studies, has allowed us to identify the structure and factors associated to HIV-1 strains whose transmission is established in Spain.

TCs were found associated with male gender and MSM. In fact, almost all the largest TCs (≥30 patients) identified in our study were associated with MSM. HIV-1 molecular epidemiological studies in different countries, independently of the methodology used, have found a similar association (Duran-Ramirez et al., 2021; Parczewski et al., 2017; Wertheim et al., 2017). Also, studies in other Spanish regions have found this association of TCs with MSM (González-Domenech et al., 2020; Pérez-Parra et al., 2015). These findings indicate that MSM TCs are playing a main role in spreading HIV-1 infections and are consistent with the fact that MSM is the main frequent mode of HIV-1 transmission in Spain since the early 2000 (Nuñez et al., 2018).

Spaniards showed higher odds than Latin Americans or Africans of belonging to TCs. However, there were important differences between these migrant populations, as 35% of Latin Americans were included in TCs, compared to only 1.2% of Sub-Saharan Africans, suggesting that HIV-1 transmission between Latin Americans and Spaniards is relatively common, presumably derived from cultural and linguistic affinities. Similarly to our results, a low percentage of Sub-Saharan Africans in TCs have been also reported in Italy (Fabeni et al., 2020) and Belgium (Verhofstede et al., 2018). In our study, TCs are mainly MSM-driven. In contrast, we have found high frequencies of females (52%) and heterosexuals (88%) in Sub-Saharan Africans, which, in addition to the likely HIV-1 acquisition in their countries of origin, can explain the low proportion of individuals in this migrant population belonging to TCs. Indeed, we have observed that Sub-Saharan Africans are infected frequently with genetic forms circulating in Africa.

In Spain, 36% of HIV-1 infections diagnosed in 2019 were in migrants (HIV, STI and hepatitis surveillance unit, 2020). Determining the origin of HIV-1 infections in the migrant population would allow proper allocation of resources for preventive measures (Álvarez del Arco et al., 2017; Fakoya et al., 2015). A previous study based on models using clinical and behavioral data found a high level of post-migration HIV acquisition, up to 71% of the migrants in Spain (Álvarez del Arco et al., 2017). However, phylogenetic analyses can provide additional information on the geographical area of HIV-1 acquisition, decreasing the bias due to data collection or failure to consider the infections acquired by migrants when visiting their countries (Álvarez del Arco et al., 2017), as migrants’ mobility was associated with increased risk for HIV acquisition (Dias et al., 2020). In our study, the lower percentage of foreigners found in TCs could be due to infections with virus strains which are not circulating in Spain, suggestive of imported infections, which would be especially frequent in Sub–Saharan Africans. Further studies using a combined phylogenetic and epidemiological approach can address the accuracy of the origin of the HIV-1 infections in the migrant populations in Spain.

Subtype B was the most frequent genetic form associated to TCs. Only subtype F infections (76% of NDs in TCs) showed a higher probability than subtype B infections (56% of NDs in TCs) of belonging to TCs. This was mainly due to the presence of two large TCs of F1 subsubtype, designated F1_1 (205 individuals) and F1_3 (36 individuals), which have spread among MSM mainly in Galicia and Basque Country, respectively (Delgado et al., 2015; Delgado et al., 2019). Interestingly, two other large non-subtype B TCs associated with MSM, of CRF02_AG and A1 subsubtype, are also present in these regions (Delgado et al., 2019). In Western Europe, the HIV-1 epidemic among MSM is dominated by subtype B. However, the epidemic among MSM in Spain is becoming increasingly diverse through the expansion of multiple non-subtype B TCs, comprising or related to viruses circulating in other countries (Delgado et al., 2019). This phenomenon has been also documented in other European countries (Dauwe et al., 2015; Duran Ramirez et al., 2021; Fabeni et al., 2019; Ragonnet-Cronin et al., 2016; Verhofstede et al., 2018), where viruses introduced from abroad have expanded successfully among the MSM population.

A different distribution of TCs is responsible for the spread of HIV-1 in Galicia and the Basque Country. Large MSM TCs have spread locally in both regions and the majority of the identified TCs were associated with this population group. However, we found a higher frequency of TCs associated with PWID or heterosexual transmission in Galicia. Galician patients have 1.5 higher odds of belonging to TCs than individuals from the Basque Country. This can be related to the low percentage of migrants in Galicia, who have a reduced association to TCs, and the high number of patients belonging to the large F1_1 TC, currently comprising 205 individuals, which has successfully spread in Galicia and other Spanish regions (Delgado et al., 2015), even in other European countries (Delgado et al., 2015; Vinken et al., 2019). We have found an association of young patients (age range 20-29 years) to TCs in Galicia, similarly to the findings in other studies (Brenner et al., 2017; Dennis et al., 2018; Fabeni et al., 2019; González-Domenech et al., 2020; Paraskevis et al., 2019; Verhofstede et al., 2018), but not in the Basque Country. Public health interventions for reducing high risk behavior and control measures in this age group in Galicia should be implemented to decrease HIV-1 transmissions in the region. Moreover, our results highlight that even regions with comparable demographic features can show a different HIV-1 epidemic pattern, likely due to specific social and cultural differences at regional level.

In this study we have shown that MSM TCs are a keystone of the HIV-1 epidemic at regional and country levels. Consequently, reinforcement of public health measures for preventing HIV-1 infections in MSM, such as behavioral interventions to reduce risky practices, pre-exposure prophylaxis (Grant et al., 2010; McCormack et al., 2016; Riddell et al., 2018), and early diagnosis and treatment of HIV-1 infections (European Centre for Disease Prevention and Control, 2015; United Nations Population Fund, Global Forum on MSM & HIV, United Nations Development Programme, World Health Organization, United States Agency for International Developmenty, World Bank, 2015) are recommended. In addition, new strategies and other tools for reaching the MSM population, such as the apps for sex dating suggested in other studies (Duncan et al., 2018; Wirden et al., 2019) could be considered for preventing HIV-1 infections.

Implementation of measures targeting MSM could reduce indirectly the spread of HIV-1, which is mainly MSM-driven, in other population groups. National surveillance data in the USA suggest that infections among heterosexual women predominantly originate from MSM (Oster et al., 2015). Moreover, the presence of female and self-declared heterosexual male individuals in MSM TCs is frequent (Esbjörnsson et al., 2016; Hué et al., 2014; Ragonnet-Cronin et al., 2018; Verhofstede et al., 2018), as we have also observed in large TCs in Spain (Delgado et al., 2019). Analyses to evaluate the contribution of MSM in HIV-1 transmission to other population groups are needed in Spain.

The high representativeness of our cohort, especially in the Basque Country, suggests that the largest and most spread TCs in these regions have been identified, which supports the conclusions of the study. However, our data may have some limitations as some of the NDs from Basque Country and Galicia during the studied period were not analyzed and not all the TCs propagating in Spain were detected in our phylogenetic analysis.

Our study has provided valuable data on the HIV-1 epidemic in Spain, identifying specific features at geographical and population levels for tailoring more specific public health interventions. New approaches and strategies are needed to reach the UNAIDS 90-90-90 target proposed by 2020 (UNAIDS, 2014). To achieve this goal, the implementation of a molecular HIV-1 surveillance system to monitor the evolution of the epidemic in Spain could be of great help. Such a surveillance system could allow to promptly detect rapidly expanding TCs needing urgent investigation and the implementation of additional public health measures to prevent their spread (Centers for Disease Control and Prevention, 2018), as well as to readjust the control measures in place according to the evolution of the TCs.

## Data Availability

The raw data supporting the conclusions of this article will be made available by the authors, without undue
reservation.
In

## MEMBERS OF THE SPANISH GROUP FOR THE STUDY OF NEW HIV DIAGNOSES

**Basque Country:** Hospital Universitario de Araba, Vitoria: Andrés Canut-Blasco, José Joaquín Portu, Carmen Gómez-González; Hospital Universitario de Basurto, Bilbao: Josefa Muñoz, M<u>^a^</u> Carmen Nieto, María Zuriñe Zubero, Silvia Hernáez-Crespo, Estibaliz Ugalde; Hospital Universitario de Cruces, Barakaldo: Luis Elorduy, Elena Bereciartua, Leyre López Soria; Hospital de Galdakao: M<u>^a^</u> José López de Goicoechea, José Mayo; Hospital Universitario Donostia: Gustavo Cilla, Julio Arrizabalaga, José Antonio Iribarren, M<u>^a^</u> Jesús Bustinduy, M<u>^a^</u> Julia Echevarría. M<u>^a^</u> Yolanda Salicio, David Grandioso. **Galicia:** Área sanitaria de Ferrol: Ana Mariño, Patricia Ordóñez, Hortensia Álvarez, Nieves Valcarce; Complejo Hospitalario Universitario de A Coruña: Ángeles Cañizares, M<u>^a^</u> Ángeles Castro. Hospital Universitario Lucus Augusti, Lugo: Ramón Rabuñal-Rey, María José García-País, M<u>^a^</u> José Gude-González, Pilar Alonso-García, Antonio Moreno-Flores; Complejo Hospitalario Universitario de Ourense: Juan García Costa, Ricardo Fernández-Rodríguez, Raúl Rodríguez-Pérez, Jorge Guitián, María Dolores Díaz-López, María Genoveva Naval-Calviño; Complejo Hospitalario Universitario de Vigo: Celia Miralles, Antonio Ocampo, Sonia Pérez-Castro, Jorge Julio Cabrera; Complejo Hospitalario Universitario de Pontevedra: Julio Díz-Arén, Matilde Trigo, M<u>^a^</u> Ángeles Pallarés. **Navarra:** Complejo Hospitalario de Navarra, Pamplona: Carmen Ezpeleta Baquedano, Carmen Martín Salas, Irati Arregui García, María Gracia Ruiz de Alda. **Madrid:** Centro Sanitario Sandoval, Madrid: Jorge del Romero, Carmen Rodríguez, Mar Vera, Óskar Ayerdi, Eva Orviz; Hospital Universitario de Fuenlabrada: María Isabel García-Arata, Santiago Prieto-Menchero; Hospital Clínico Universitario San Carlos, Madrid: Esther Culebras, Icíar Rodríguez-Avial; Hospital Universitario Fundación Jiménez Díaz, Madrid: Raquel Téllez-Pérez, Olalla Calabia-González, Alfonso Cabello-Úbeda, Miguel Górgolas Hernández-Mora; Hospital Universitario Severo Ochoa, Leganés: Sara María Quevedo, Lucía Puente, Manuel del Álamo; Hospital Fundación Alcorcón, Madrid: Carolina Campelo Gutiérrez, María José Goyanes Galán; **Castilla y León:** Hospital Clínico Universitario de Valladolid: Carmen Hinojosa, Carlos Dueñas, Begoña Monteagudo, Edita Sánchez; Hospital Río Hortega, Valladolid: Jessica Abadía, Belén Lorenzo Vidal; Hospital Virgen de la Concha, Zamora: Teresa Martín-Domínguez, Rosa Martínez-González. **La Rioja:** Hospital San Pedro, Logroño: José Ramón Blanco, Miriam Blasco. **Aragón:** Hospital Universitario Miguel Servet, Zaragoza: Ana María Martínez-Sapiña, Piedad Arazo. **Castilla-La Mancha**: Hospital Universitario de Guadalajara: Alejandro González Praetorius, Complejo Hospitalario de Toledo: César Gómez-Hernando, José Largo-Pau; **Comunitat Valenciana:** Hospital Universitari Sant Joan d’Alacant: Fernando Buñuel, Ana Infante.

## ETHICS STATEMENT

Sequences were derived from antiretroviral resistance genotypic test. The use of anonymized, de-identified clinical/demographic and sequence data was reviewed and approved under an exempt protocol by the Bioethics and Animal Well-being Committee of Instituto de Salud Carlos III, with report numbers CEI PI 38_2016-v3 (dated 20 June 2016) and CEI PI 31_2019-v5 (dated 6 November 2019). This study did not require written informed consent by study participants, except for those participants who required additional samples different from the ones obtained in the routine clinical practice.

## AUTHOR CONTRIBUTIONS

HG, ED, and MT conceived the study and supervised the experimental work. ED, HG, MT, and JC processed sequences and performed phylogenetic analyses. HG and ED performed data curation. HG, AD, and LG performed statistical analyses. SB, MS, VM and EG-B performed experimental work. The members of the Spanish Group for the Study of New HIV Diagnoses recruited patients and obtained epidemiological and clinical data. HG wrote the manuscript draft, with contributions to the text by AD, MT and ED. All authors read and approved the text.

## ACKNOWLEDGEMENTS

We would like to thank José Antonio Taboada, from Consellería de Sanidade, Xunta de Galicia, and Daniel Zulaika, from Osakidetza-Servicio Vasco de Salud, for their support of this study, and the personnel at the Genomic Unit, Centro Nacional de Microbiología, Instituto de Salud Carlos III, for technical assistance in sequencing.

