## Supplementary tables for "Transmission clusters, predominantly associated with men who have sex with men, play a main role in the propagation of HIV-1 in Northern Spain (2013-2018)"

**Supplementary table 1:** Reagents and condition of the Pr-RT amplification.

| Reagents | RT-PCR  (for RNA) | 1^st^ PCR  (for DNA) | Nested PCR  (RNA and DNA) |
| --- | --- | --- | --- |
| OneStep RT-PCR (QIAGEN) enzyme | 1 ul | -- | -- |
| GoTaq G2 Hot Start colorless mastermix (Promega) | -- | 12.5 ul | -- |
| BioTaq DNA polymerase (Bioline) | -- | -- | 0.75 U |
| RNAse inhibitor | 5 U | -- | - |
| MgCl_2_ | 2.5 mM* | 2mM† | 1.8 mM |
| dNTPs | 400 µM | 200 µM† | 200 µM |
| Each Primer | 0,6 µM | 0,4 µM | 0,4 µM |
| Reaction volume | 25 µl | 25 µl | 25 µl |
| Thermal cycling profile |  |  |  |
| Reverse transcription | 30 min 50ºC | **--** | **--** |
| Hot start activation | 15 min 95ºC | **--** | **--** |
| Denaturation | -- | 2 min 94ºC | 2 min 94ºC |
| Cycling | 40 | 35 | 40 |
| Denaturation | 30 sec 94ºC | 30 sec 94ºC | 30 sec 94ºC |
| Annealing | 1 min 55ºC | 1 min 55ºC | 1 min 56ºC |
| Extension | 90 sec 72ºC | 90 sec 72ºC | 1 min 72ºC |
| Final extension | 10 min 72ºC | 7 min 72ºC | 5 min 72ºC |

***** included in the 2x kit mastermix; † Included in the 5x kit buffer.

**Supplementary table 2:** Primers used in the Pr-RT amplification and sequencing.

| Oligo | Use | Direction | HXB2 position | Sequence |
| --- | --- | --- | --- | --- |
| RP-1-S | 1st PCR/RT-PCR | sense | 2016-2041 | 5’-GAA AAA GGG CTG TTG GAA ATG TGG AA |
| RP-1-A | 1st PCR/RT-PCR | antisense | 3685-3716 | 5’-AAA TTT AGG AGT CTT TCC CCA TAT TAC TAT GC |
| PR-O-S2b | nested PCR/ sequencing | sense | 2080-2107 | 5’-GCT AAT TTT TTA GGG AAR ATY TGG CCT T |
| RT-O-A | nested PCR/ sequencing | antisense | 3630-3662 | 5’-TGC CTC TGT TAA TTG TTT TAC ATC ATT AGT GTG |
| PRsec2A | sequencing | sense | 2838-2811 | 5’-GAT GYG GTA TTC CTA ATT GRA CYT CCC A |
| RTsec1S | sequencing | antisense | 2692-2716 | 5´-CAA AAA TTG GGC CTGA AAA TCCA TA |
